# Global Determinants of Covid-19 Deaths: Lockdown Dates and Social Distancing Measures Mattered

**DOI:** 10.1101/2020.07.28.20163394

**Authors:** Vince Hooper

## Abstract

**Objectives:** The objective of this paper is to examine the influence that various contextual variables have upon the number of deaths due to covid-19, across the world.

**Setting Level:** This study utilizes data for 125 countries for contextual variables from 1st January 2020 until the 15th June 2020.

**Participants:** This study considers deaths from covid-19.

**Interventions:** DELETED

**Primary and secondary outcome measures:** The contextual variables considered in this study are stringency index, stringency variability, lockdown date, population density, level of airline passengers and country health security index.

**Results:** It is shown there is a very strong association between the level of airline passengers and covid-19 deaths. The results from regression analysis conducted in this study show significant positive relationships at the 5% level of statistical significance between Deaths from covid-19 and airline passenger levels and stringency variability; significant negative relationships are revealed for stringency index and lockdown date supporting the notion that lock down and social distancing measures mattered and were effective. The Global health security index and population density did not significantly affect deaths.

**Conclusion:** This study highlights the strong link between a country’s airline passengers and covid-19 deaths and found that the lockdown date and stringency measures had a significant effect upon deaths. The implications of the research is that lockdown and stringency measures implemented by governments around the world worked and mattered. Further, the fact that global health security did not affect deaths may indicate better preparedness required to confront future pandemics.

**Trial Registration:** DELETED

1. FUNDING: This research received no specific grant from any funding agency in the public, commercial or not-for-profit sectors.

**Article Summary: Strengths and Limitations of Study:**

- It is discovered in this paper, for a sample of 125 countries that lockdown and social distancing measures had a very significant positive effect upon reducing covid-19 deaths across the world. Countries deaths were very significantly positively related to the level of annual airline passengers. A combination of 18 countries with a share of 84% of global annual air passengers accounted for 80% of total deaths recorded from covid-19, worldwide.
- The quality of a country’s health system as measured by a new measure GHI Global Health Security Index did not significantly reduce the number of covid-19 deaths, supporting the fact that both developed and developing countries were lacking in essential equipment, as well as track and trace mechanisms.
- The strengths of the study are that it is very timely and judgement needs to be made at the national and international level whether the lockdown was effective given the likelihood of a 2^nd^ wave to the pandemic. Countries need to better prepare themselves for future pandemics in terms of rapid data sharing and analysis to ensure that outbreaks are contained more effectively and efficiently through stringent lockdown and social distancing measures.
- A limitation of the study is the quality of data relating to the deaths actually caused by the covid-19 virus. Despite this the methodology and results of the paper are very sound and robust.

## 1. Introduction

The severity of this global pandemic and focus upon its devastating effects upon local communities has tended to be through the media with time series graphics at the national level. Thus, the purpose of this paper is to examine the cross-national differences in the deaths caused by covid-19, and relate that to a range of contextual variables.

The covid-19 pandemic is unprecedented in modern times and there is a scarcity of knowledge at this present time in relation to the effectiveness of lockdown and social distancing measures. However, there is evidence to suggest, on a limited country time series basis that lockdown measures flattened the initial wave of the pandemic when they were eventually implemented. It has been argued that the WHO World Health Organization was slow in declaring covid-19, a global pandemic. Subsequently, national governments were late in their response to the pandemic. Countries were ill prepared to deal with this pandemic in terms of quality data collection and analysis; the availability of ventilators as well as personal protection equipment.

The sample considered in this paper is 125 countries over the time period 1st January to 15th June 2020. The contextual variables are the stringency index, stringency variability, lockdown date, population density, level of airline passengers, country health security index and whether the lockdown was full, partial or none.

Whilst it is recognized that data sources need to be improved, validated and verified, the results of this paper are quite robust at this stage in our understanding of covid-19, in general.

A very high association is found between annual airline passengers and covid-19 deaths. In particular countries that have the highest share of airline passengers internally, outward and inward bound have the highest deaths in the world consistent with the view that viruses become internationalized rapidly through rapid travel flows across international frontiers via airliners.

The paper has important policy implications in the sense that we need to monitor more accurately international travelers’ health, in real time and contact with others more effectively and efficiently than previously, perhaps through a ‘test, track and trace’ type app on a smartphone. This has obvious implications for privacy, civil liberties and individual freedoms which need to be weighed up against the risk of spreading disease and closing the world down.

This paper is organized as follows:

Section 2, outlines the data sources. Section 3, discusses the results. Section 4 summarizes the paper and makes some public health policy suggestions. Section 5 presents the references.

## 2. Data

The population density data are sourced from the World Bank Development Indicators database [1]. Annual Airline Passengers 2019 are extracted from World Air Transport Statistics [2]. The Stringency Index is sourced from Oxford University [3], “Coronavirus Government Response Tracker”. Mean and variability (standard deviation) stringency index is computed over the study period from 1st January to 15th June 2020. Lock down dates and status is sourced from [4] & Country Health Security Index 2019 is sourced from [5]. Deaths, worldwide from covid-19 are sourced from Worldometers [6]. Data was from reliable and collected ethically by the providers and this study did not need the consent of patients directly, and is publicly available.

The period under study for deaths and lock down is 1st January 2020 until 15th June 2020.

## 3. Results

Table 1 highlights the very robust relationship between the country ranking of annual airline passengers and covd-19 deaths. At the top of the table, a combination of 18 countries account for 3.66 billion passengers, which corresponds to 84% of annual passengers carried in the world, annually. Also in Table 1, there is a corresponding figure for the number of covid-19 deaths for these 18 countries which represents 80% of overall deaths in the world up to the 15th June 2020.

**Table 1:**

| **Country** | **Airline**<br>**Passengers** | **Covid-19**<br>**Deaths** |
| --- | --- | --- |
| United States | 796877823 | 121441 |
| China | 668024219 | 4634 |
| United Kingdom | 251370641 | 42589 |
| Spain | 200814594 | 28322 |
| Japan | 187233922 | 952 |
| India | 176719682 | 13254 |
| Germany | 171126916 | 8961 |
| Italy | 146984636 | 34610 |
| France | 140165675 | 29633 |
| Indonesia | 138807314 | 2429 |
| Thailand | 105905344 | 58 |
| Korea (Republic) | 102304402 | 280 |
| Russian | 100491546 | 8002 |
| Canada | 97545561 | 8410 |
| Australia | 97486223 | 102 |
| Turkey | 95792929 | 4927 |
| Brazil | 93266806 | 50058 |
| Mexico | 88900965 | 20781 |
| Sum of Table<br>Column | 3659819198 | 379443 |
| Total Airline<br>Passengers | 4377670000 | 472000 |
|  | 84% | 80% |

Indeed when a rank correlation coefficient is computed between number airline passengers and death for 126 countries, this gives an association r=0.74, at a 1% significance level. Table 2 reveals the results of the regression in analysis:

**Table 2:** Regression Model Summary.

| Model | R | R Square | Adjusted R Square | Std. Error of the Estimate |
| --- | --- | --- | --- | --- |
| 1 | .856 | .733 | .716 | 7251.943 |

Coefficients
| Model | Independent Variables | Unstandardized Coefficients |  | Standardized Coefficients | t | Sig. |
| --- | --- | --- | --- | --- | --- | --- |
|  |  | B | Std. Error | Beta |  |  |
| 1 | (Constant) | -43683.0 | 10571.87 |  | -4.132 | .000 |
|  | AIRPLINE PASSENGERS | .000114 | .000008 | .915 | 13.915 | .000 |
|  | HEALTH INDEX | 110.637 | 66.059 | .106 | 1.675 | .097 |
|  | STRINGENCY INDEX | -445.201 | 165.996 | -.204 | -2.682 | .009 |
|  | STRINGENCY VARIABILITY | 745.247 | 196.946 | .304 | 3.784 | .000 |
|  | POPULATION DENSITY | .112 | .363 | .017 | .310 | .757 |
|  | LOCK DOWN DAY | 382.334 | 95.146 | .247 | 4.018 | .000 |

Deaths= constant + ß1 Airline Passengers + ß2 Health Index + ß3 Stringency Index + ß4 Stringency Variability + ß5 Population Density + ß6 Lockdown Day + Error.

The Dependent Variable was the NUMBER OF DEATHS for 125 COUNTRIES. Residual errors are normally distributed, as supported by a QQ Plot, in the regression model.

The regression equation results in a high R-squared of order 0.70. Attention to the coefficients ß1 to ß6 reveals that Health Index is significant at the 10% level and Population Density is insignificant. Health service issues were at the centre of major issues confronting all countries given the scarcity of ventilators and personal protection equipment. It appears other issues were highly significant. Deaths are positively significantly (at the 1% level) in relation to Airline Passengers and Stringency Variability; and inversely significantly related to Stringency Index and Lockdown date.

This regression thus shows that the ‘global’ lockdown had a significant effect upon reducing deaths. More specifically, each day of extra lockdown, from the regression equation, would have saved approximately 400 lives per country.

## 4. Summary and Health Policy Recommendations

It is exposed in this paper the very strong link between air travel and the spread of the pandemic. However, despite this finding it needs to be stressed that the control of this pandemic has been to use both domestic and international policy lockdown tools, in conjunction with social distancing measures. The regression analysis in this paper deems these policy tools to be highly effective in reducing deaths from covid-19. Despite air travel having greater security checks since 911, a possible unfortunate yet necessary step may be to monitor travelers’ health more rigorously through the use of mobile phone track and trace technology, across international frontiers coupled with greater testing capabilities. Health index producers and the WHO World Health Organization may also look closely at different nations’ responses to the covid-19 and incorporate this as an important parameter in their future data collection, to enhance their explanatory power. This may include covering and reviewing basic items like personal protection equipment for hospital staff.

## Data Availability

Data is publicly available.

## FURTHER POINTS ADDRESSED

**‘Patient and Public Involvement’ [none involved]**

[My responses in italics].

- How was the development of the research question and outcome measures informed by patients’ priorities, experience, and preferences?

*Data on covid-19 deaths, worldwide, was collected from a publicly available database from Worldometers*..

*[https://www.worldometers.info/coronavirus/]. “Reported Cases and Deaths by Country, Territory, or Conveyance”*.

*“Worldometer manually analyzes, validates, and aggregates data from thousands of sources in real time and provides global* ***COVID-19 live statistics*** *for a* ***wide audience of caring people around the world***.

***Our data is also trusted and used by*** *the [UK Government](https://assets.publishing.service.gov.uk/government/uploads/system/uploads/attachment_data/file/878126/COVID-19_Press_Conference_Slides_05042020.pdf), [Johns Hopkins CSSE](https://gisanddata.maps.arcgis.com/apps/opsdashboard/index.html#/bda7594740fd40299423467b48e9ecf6), the [Government of Thailand](https://ddc.moph.go.th/viralpneumonia/eng/index.php), the [Government of Vietnam](https://ncov.moh.gov.vn/), the [Government of Pakistan](http://covid.gov.pk/stats/global), [Financial Times](https://www.ft.com/coronavirus-latest), [The New York Times](https://www.nytimes.com/2020/03/20/health/coronavirus-data-logarithm-chart.html), [Business Insider](https://www.businessinsider.com/figures-show-us-soon-coronavirus-worse-china-2020-3), [BBC](https://www.bbc.co.uk/iplayer/episode/m000h499/bbc-news-at-ten-09042020), and many others*.

*Over the past 15 years, our statistics have been requested by, and provided to* ***Oxford University Press, Wiley, Pearson, CERN, World Wide Web Consortium (W3C), The Atlantic, BBC***, *Milton J. Rubenstein Museum of Science & Technology, Science Museum of Virginia*, ***Morgan Stanley, IBM, Hewlett Packard, Dell, Kaspersky, PricewaterhouseCoopers, Amazon Alexa, Google Translate***, *the* ***United Nations Conference on Sustainable Development (Rio+20)***, *the [U2 concert](https://www.worldometers.info/u2/), and many others*.

*Worldometer is cited as a source in over 10,000 published books and in more than [6,000 professional journal articles](http://scholar.google.com/scholar?hl=en&q=worldometers&btnG=&as_sdt=1%2C22&as_sdtp=) and was voted as one of the [best free reference websites](http://www.ala.org/rusa/sections/mars/marspubs/marsbestfreewebsites/marsbestref2011) by the* ***American Library Association*** *(ALA), the oldest and largest library association in the world.”*

- How did you involve patients in the design of this study? *Not relevant*
- Were patients involved in the recruitment to and conduct of the study? *Not relevant*
- How will the results be disseminated to study participants? *Not relevant*

## Notes

### Competing Interest Statement

The authors have declared no competing interest.

